# Genome-Wide Analysis Reveals a Role of 3’-untranslated Region Variants Affecting Cleavage and Polyadenylation in Undiagnosed Rare Disorders

**DOI:** 10.1101/2025.02.16.25322367

**Authors:** Y. Kainov, C. Dias, T. Hubbard

## Abstract

Cleavage and polyadenylation of pre-mRNAs are essential for transcription termination and the normal expression of eukaryotic genes. However, the extent to which mutations in cleavage and polyadenylation signals act as causal variants in rare Mendelian disorders remains unknown. Using deep learning models, we identified enrichment of alleles predicted to disrupt polyadenylation in a large cohort of undiagnosed probands with rare disease disorders from the Genomics England 100kGP dataset. These alleles were predominantly located in the 3’ UTRs of genes known to be causally associated with rare diseases. Using the *SLC16A2* gene linked to Allan-Herndon-Dudley syndrome as an example, we show that a putative causal variant predicted to disrupt cleavage and polyadenylation in a proband with intellectual disability has a deleterious impact on *SLC16A2* expression. Our findings highlight the underappreciated contribution of cleavage and polyadenylation-disrupting variants to the genetic basis of rare diseases.

## Introduction

The maturation of eukaryotic mRNAs involves several essential steps, including the addition of a 5’ 7-methylguanosine cap, intron removal through splicing, and cleavage followed by polyadenylation at 3’ end. The latter plays a crucial role in transcription termination and the release of RNA transcripts from elongating RNA polymerase II, ensuring productive gene expression (Rodríguez-Molina *et al*, 2023; Mitschka & Mayr, 2022). Mechanistically, cleavage and polyadenylation require the co-transcriptional assembly of multisubunit protein complex, driven by various polyadenylation signals (PAS), with AWTAAA (and its variants, commonly referred to as PAS hexamers) being the strongest and most prevalent (Proudfoot, 2011).

PAS hexamers and other polyadenylation signals are subject to purifying selection (Kainov *et al*, 2016; Findlay *et al*, 2024; Kainov *et al*, 2024) with disruption by a limited known number of germline variants associated with heightened cancer susceptibility and rare genetic syndromes (Johnston *et al*, 2019; Bennett *et al*, 2001; Higgs *et al*, 1983; Stacey *et al*, 2011; Xiao *et al*, 2023). RNA-seq-centered studies have only recently begun to elucidate the impact of genetic variants affecting alternative polyadenylation on complex diseases and traits (Li *et al*, 2021; Zou *et al*, 2025).

In case of rare diseases, previous systematic efforts mostly focusing on non-coding variants in general have not detected novel causative pathogenic variants near polyadenylation sites, likely because of limitations of used variant effect prediction approaches, rare nature of the variants, and small dataset sizes (Linder *et al*, 2022; Martin-Geary *et al*, 2023; Lord *et al*, 2024).

Here, starting with variant effect predictions from advanced machine learning models, we conducted a systematic genome-wide analysis of germline variants inferred to impact cleavage and polyadenylation in mRNA 3’-untranslated regions (3’UTRs) in individuals with rare diseases in the Genomics England (GEL) 100,000 Genomes Project (100kGP) dataset (National Genomic Research Library, 2017). We detected a significant enrichment of rare variants predicted to suppress polyadenylation in undiagnosed rare disease patients compared to diagnosed patients and provided further experimental evidence of the disruptive effect of such variants on corresponding gene expression.

## Results

### Genome-wide identification of rare variants disrupting polyadenylation (DOWNvars)

To understand the potential role of variants affecting polyadenylation in undiagnosed rare diseases, we annotated all short variants (single-nucleotide variants (SNVs) and insertions and deletions (InDels) ≤50 bp) near cleavage and polyadenylation sites (CS) located in the 3’ UTRs of coding genes using the PolyaID and PolyaStrength deep-learning models. (Stroup & Ji, 2023). Across genome-wide variant profiles of 24,246 probands with the clinical manifestations of a suspected Mendelian disorder, we identified all variants for which there was at least one allele present across the cohorts that were predicted to strongly downregulate polyadenylation at the given strong CS - DOWNvars (see Methods for details).

As expected, DOWNvars were clustered immediately upstream of annotated CSs where the major polyadenylation signal (PAS-hexamer with AWTAAA consensus) is located (Fig1A). Additionally, we observed a smaller peak downstream of CS, likely representing disruptions of GU-rich downstream elements (Nunes *et al*, 2010) (Fig1A). Consistent with the observed positional pattern we identified a striking enrichment of DOWNvars disrupting the two strongest PAS-hexamer variants (AATAAA and ATTAAA) in comparison to predicted non-disruptive variants in the same regions (‘background variants’, see Methods, 43.35% of events vs 0.84% of events, two-tailed Fisher test P=0). Additionally, DOWNvars were more likely to be deletions (Fig1B).

**Figure 1.**
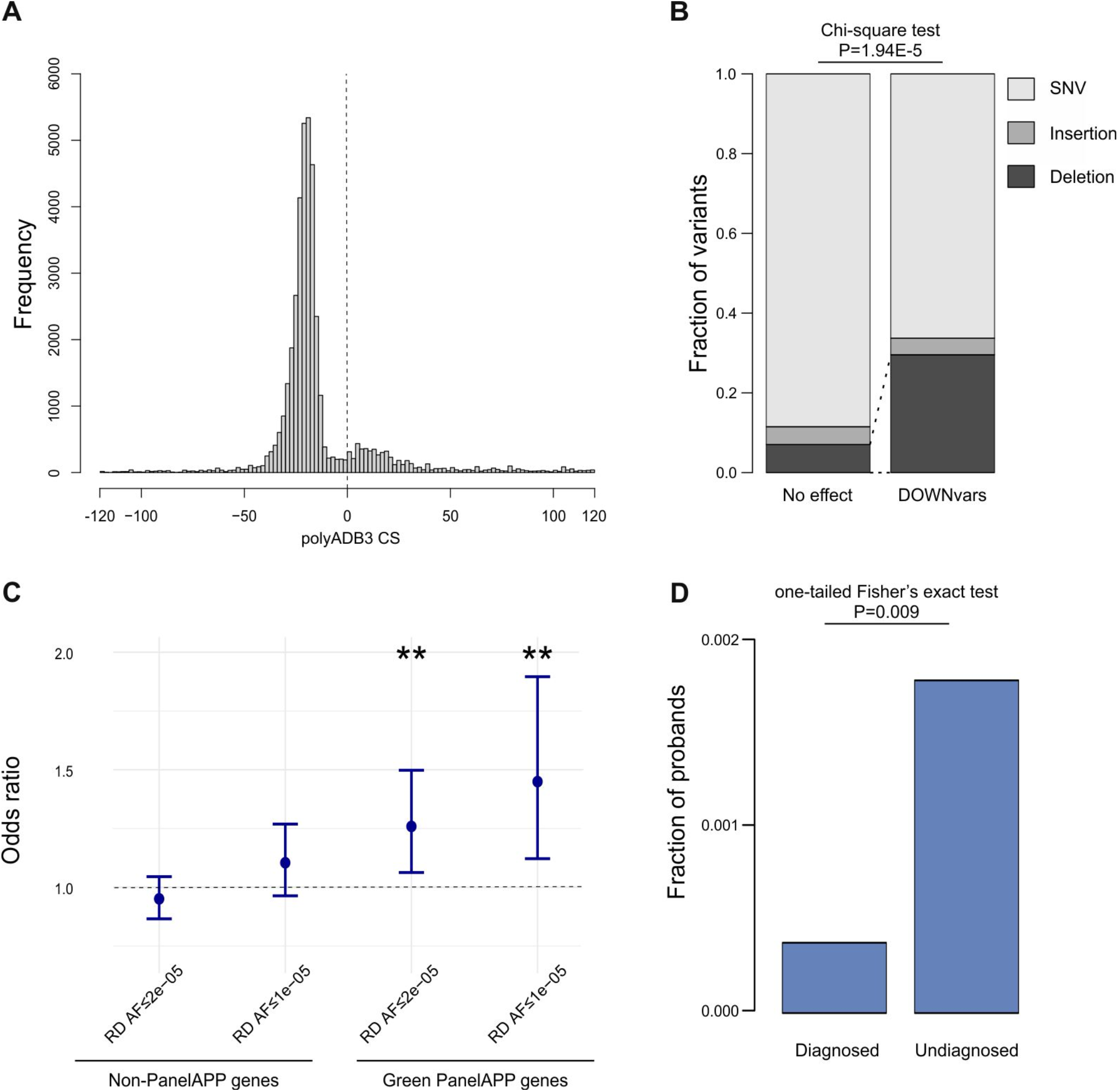
**A** – Positional distribution of DOWNvars in relation to cleavage and polyadenylation site (CS), note strong enrichment 20-25 bp upstream of CS (position of PAS hexamer) and moderate enrichment immediately downstream of CS (position of GU-rich tract). **B** - Bar plot showing enrichment of deletions in DOWNvars subset. **C** - Odds ratios for an enrichment of ultrarare DOWNvars (not gnomAD, GEL AF≤0.00002) in undiagnosed probands in comparison with diagnosed probands in green PanelApp genes (right) and other genes (left), whiskers represent 95% confidence intervals. ** - BH-adjusted one-tailed Fisher’s exact test p-value < 0.01. **D** – Bar plot showing enrichment of ultrarare DOWNvars (not gnomAD, GEL AF≤0.00001) in undiagnosed probands within dominant green PanelApp genes when the proband’s normalized disease group was matched to the corresponding PanelApp gene.

### Rare DOWNvars are enriched in undiagnosed probands in genes implicated in rare diseases

To evaluate if this set of DOWNvars is enriched for potential causal alleles we compared the frequency of DOWNvars alleles observed in undiagnosed probands to the frequency observed in diagnosed probands (see Methods for details and Table S1). Whilst no enrichment was observed across all DOWNvars (FigS1), stratification by allele frequency revealed evidence of significant enrichment of DOWNvars alleles with the lowest population frequency (variants unobserved in gnomAD and singletons in GEL RD cohort, BH-adjusted P=0.007, FigS2).

We hypothesised that if ultrarare DOWNvars are enriched for pathogenic variants, these variants would be significantly overrepresented in the cleavage and polyadenylation sites of genes where protein-coding variants have previously been implicated as the causes of rare disease. To test this, we utilised the set of genes with confirmed Mendelian disease association from the Genomics England’s PanelApp (Stark *et al*, 2021). Using this set of 4091 diagnostic-grade genes with definitive evidence for a causal role in disease, we observed a strong enrichment of ultrarare DOWNvars alleles, but not in genes without known links to rare human diseases (green PanelApp genes) (Fig1C). This observation was prominent in both twice recurring (two alleles per GEL RD dataset) and unique (one variant per GEL RD dataset) DOWNvars. Specifically, these ultrarare DOWNvars were significantly enriched in dominant green PanelApp genes (monoallelic mode of inheritance) in line with their extremely low allele frequencies and likely disrupting nature (FigS3).

Notably, the strength of this observed enrichment of unique variants increased when the undiagnosed probands’ phenotype was matched to the corresponding PanelApp gene sets (“Normalized disease group” for proband’s phenotype in GEL RD being identical to the “Panel Disease Group” for corresponding PanelApp gene, Fig1D, OR=4.75, OR CI=1.22-40.79, P=0.0087). These results suggest that rare variants affecting polyadenylation might play a critical role in pathogenesis of rare diseases in a subset of undiagnosed probands.

Our findings were reproducible separately in probands of genetically inferred European ancestry and probands of other ancestries (FigS4). Interestingly, we did not observe significant enrichment of variants affecting polyadenylation in weaker CS (PolyaID predicted probability of being a CS<0.9) in green PanelApp genes (FigS5). These results suggest that the observed burden of DOWNvars in undiagnosed probands is unlikely to be due to the non-specific enrichment of rare variants from underrepresented genetic ancestries (Tallman *et al*, 2024) or differences in local susceptibility to mutation.

### DOWNvar in causative gene SLC16A2 impairs corresponding gene expression

To identify variants with potential clinical significance, we manually compared the Human Phenotype Ontology (HPO) terms recorded at the time of proband recruitment to the phenotypes typically associated with heterozygous loss-of-function or hypomorphic variants in the corresponding gene, assuming that DOWNvars are likely to reduce gene expression (Kainov *et al*, 2024; Grzechnik & Mischo, 2025; Higgs *et al*, 1983). Specifically, unique and recurrent (twice-occurring) DOWNvars in Dominant and X-linked PanelApp genes were shortlisted based on the undiagnosed proband’s normalized disease group and subgroup. HPO terms were then manually compared with OMIM phenotypes for the corresponding conditions. Additionally, we assessed the redundancy of affected cleavage sites (presence of strong alternative polyadenylation sites), the position of the affected site within the 3’UTR, and the overall 3’UTR length.

As a result of this analysis, we selected an SNV in the terminal polyadenylation signal of the green PanelApp gene SLC16A2 (also known as monocarboxylate transporter-8 or MCT8) - chrX:74533900:A>G (GRCh38) - for further experimental validation. This variant disrupts a strong PAS hexamer (AATAAA to AATAGA), predicted to drastically decrease the polyadenylation activity at the site - polyAID (probability of being a CS) decreases from 0.99 to 0.03 (Fig 2A). The chrX:74533900:A>G is absent from gnomAD and, within the 100kGP dataset, is observed only in the proband (hemizygous) and their mother (heterozygous). Hypomorphic missense and inactivating variants in *SLC16A2* are known to cause Allan-Herndon-Dudley syndrome (Maranduba *et al*, 2006; Dumitrescu *et al*, 2004; Schwartz *et al*, 2005) (AHDS; OMIM:300523) – a condition characterised by impaired intellectual development, motor functions, and hypotonia.

**Figure 2.**
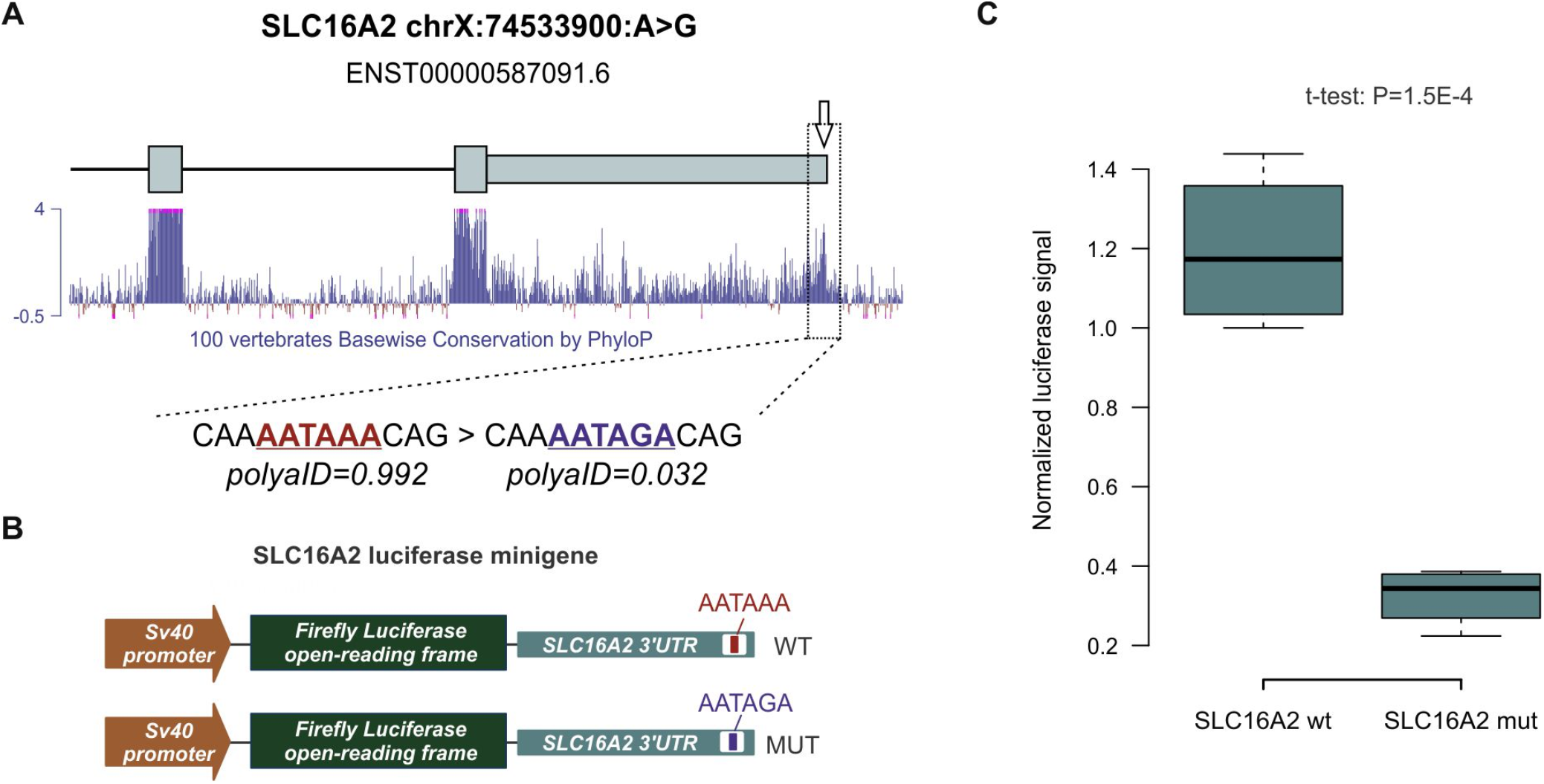
**A** – Diagram of last exons of green PanelApp gene *SLC16A2* and wild-type and mutated sequences of corresponding cleavage/polyadenylation signal (white arrow); note position of polyadenylation signal inside the peak of 100 vertebrates PhyloP conservation (blue bottom panel). **B** – Schematic representation of luciferase minigene constructs. **C** – Box plot showing that chrX:74533900:A>G variant significantly reduces the expression of the reporter gene in luciferase assay.

The hemizygous carrier of the chrX:74533900:A>G allele had been diagnosed with intellectual disability with global developmental delay and delayed gross motor development, consistent with the most common AHDS features. Additionally, the chrX:74533900:A>G variant was selected due to the known abnormal thyroid hormones levels in AHDS, which facilitate further clinical confirmation of the diagnosis, along with relatively short length of the SLC16A2 3’UTR.

To experimentally assess the effect of the chrX:74533900:A>G variant on gene expression through impairment of cleavage and polyadenylation, we cloned the 3’UTR of *SLC16A2* (chrX:74531556-74534065; GRCh38) downstream of a luciferase reporter gene, replacing original polyadenylation signals in the construct (Fig 2B). We then introduced the variant using site directed mutagenesis and analysed luciferase activity of wild type (WT) and mutated constructs in transfected HEK-293T cells. We observed a strong decrease of luciferase activity in mutant construct transfectants in comparison to wild type (Fig 2C) indicating a deleterious effect of chrX:74533900:A>G variant. Based on phenotypic match, predicted effect on cleavage and polyadenylation and an experimentally validated effect on expression, we suggest the chrX:74533900:A>G variant is Likely Pathogenic according to ACMG/AMP guidelines (Ellingford *et al*, 2022; Richards *et al*, 2015).

## Conclusion

In summary, this study underscores the pathogenic potential of ultrarare germline variants disrupting cleavage and polyadenylation in 3’UTRs in the context of rare diseases. Using a combination of deep learning predictions and rare disease-specific burden testing, we identified significant enrichment of these variants in undiagnosed probands and in genes with established disease associations. Experimental validation of a variant in *SLC16A2* further revealed the deleterious impact of polyadenylation disruption on gene expression, demonstrating potential diagnostic relevance. Our findings highlight the importance of improving predictions for the functional and pathogenic impacts of variants disrupting cleavage and polyadenylation together with their phenotypic manifestations, in order to allow their inclusion in prioritization pipelines and ultimately improve diagnostic outcomes in rare disease research.

## Methods

### Datasets and cohorts

We analysed all SNVs and InDels shorter than 51 bp in the 240bp interval centred at the 3’UTR cleavage and polyadenylation site from PolyA.db3 (Wang *et al*, 2018) in the GEL 100kGP Rare Diseases (RD) dataset Release Version 19 (for whom consent was not withdrawn; variants passed all filters in GEL aggregated vcf files - AggV2). PolyAID (Stroup & Ji, 2023) scores for reference and variant-containing sequences and corresponding differences in probability of being a site (Prob) and polyadenylation score (Score) were calculated. Sites with a Score < −0.9 were excluded from analysis. Each variant was annotated as DOWNvars (REFprob ≥0.9, VARprob <0.9 or REFprob ≥0.9, ΔScore ≤-2), DOWNweak (REFprob <0.9, ΔScore ≤-2) or background (REFprob ≥0.9, −1 < ΔScore <1). In cases where a single variant was located near more than one distinct site, the strongest effect on cleavage/polyadenylation was used for further analyses.

Participants were excluded from analysis if karyotypic and phenotypic sex was in conflict or average number of SNVs per paired and aligned number of reads were more than 2.5 Z-score or less −2.5 Z-score for that characteristic. Only variants called on genome build ‘GRCh38’ delivery version ‘V4’ were kept. Analysis was performed on unrelated individuals (individuals with a KING (Kinship-based INference for GWAS) (Manichaikul *et al*, 2010) score ≥ 0.0442 were iteratively randomly removed until no related pairs of individuals could be detected).

Analysis was performed on two cohorts – diagnosed probands (patients affected by rare diseases with established causative variant - GEL 100kGP “exit questionnaire” case flagged as “solved”, n=5279) and undiagnosed probands (patients affected by rare diseases without established causative variant - GEL 100kGP “exit questionnaire” case flagged as “no” and no diagnostic discovery was submitted, n=18967). gnomAD (v3.1.2) and GEL Rare diseases (RD) allele frequencies (AF) were obtained from AggV2 functional annotation files. Only variants with both gnomAD and RD AF ≤0.00005 were designated as rare and included in the analysis (apart from S1 where less stringent gnomAD AF thresholds were applied).

For analysis at S4 participants with a genetically inferred European origin ancestry match of 95% were designated as “European” (diag n=3801, undiag n=14565) and all other participants were combined in “other” ancestry group (diag n=1478, undiag n=4402). Table version of PanelApp containing gene-centered information was accessed and downloaded using PaneApp API and custom scripts on 18 of November 2024. For Fig1D analysis “Normalised Disease Group” field from GEL 100kGP RD dataset was compared with “Panel Disease Group” from PanelApp.

### Plasmid constructs

All plasmids were propagated in the NEB 5-alpha *E. coli* strain (NEB, cat# C2987H). To generate luciferase reporter plasmids (pYC38 and pYC39) the entire *SLC16A2* 3’UTR (chrX:74531556-74534065; GRCh38/hg38) was ordered as FragmentGENE from GENEWIZ (Azenta Life Sciences). The fragment was amplified using KAPA HiFi DNA polymerase HotStart ReadyMix (Roche, cat# KK2601) and the following primers: forward, 5’-GGAAAGATCGCCGTGTAATTACCTTTCTTGCCATTGTG-3’; and reverse, 5’-AAGGCTCTCAAGGGCATCGGTAGCTCTTTTTCTAATGTCTG-3’. The PCR product was agarose gel-purified and cloned using NEBuilder® HiFi DNA Assembly Cloning Kit (NEB, cat# E5520S) into the pGL3-Promoter plasmid (Promega, cat# U47298) after XbaI and SalI digestion. The chrX:74533900A>G variant was introduced using a modified Quikchange site-directed mutagenesis protocol, using the KAPA HiFi DNA polymerase HotStart ReadyMix (Roche, cat# KK2601) and the following primers: forward, 5’-CAAGGAGTTTGGAACAAAATAGACAGATCTAAAAATTTGG-3’ and reverse, 5’-CCAAATTTTTAGATCTGTCTATTTTGTTCCAAACTCCTTG-3’. Constructs were verified by dideoxynucleotide Sanger (Source BioScience, UK) or/and long-read sequencing (FullCirlce, UK). Plasmid and plasmid maps are available on request.

### Cell culture and luciferase assay

HEK-293T cells (ATCC, Cat# CRL-3216) were verified for morphology via light microscopy and tested negative for mycoplasma contamination using the MycoGenie Rapid Detection Kit (AssayGenie, Cat# MORV0011). Cells were maintained in DMEM (Thermo Fisher Scientific, Cat# 11360070) supplemented with 10% FBS (Hyclone, Cat# SV30160.03) at 37 °C with 5% CO_2_. For passaging, cells were washed with PBS and dissociated with 0.05% Trypsin-EDTA (Thermo Fisher Scientific, Cat# 15400054) for 10 minutes at 37 °C.

For luciferase minigene transfection experiments, cells were typically seeded overnight at 5×10^3^ in 100 μl of culture medium per well of a 96-well plate. The next morning, 70 ng of a firefly luciferase reporter construct containing wild-type and mutated 3’UTR sequences and 30 ng of the *Renilla* luciferase control (pRL-TK; Promega) were mixed with 0.1 μl of jetPRIME (Polyplus, cat#101000046) transfection reagent in 10 μl of jetPRIME transfection buffer, incubated for 10 min at RT and added dropwise to the cells. After 48 hr incubation, transfected cells were lysed and processed using a Dual-Glo Luciferase Assay System reagents (Promega, cat# E2920) as recommended by the manufacturer. Luminescence was measured using a Perkin Elmer Victor X2 plate reader.

### Statistical analysis

Unless stated otherwise, all statistical procedures were performed in R v.4.2.1. Significance of enrichment was calculated using Fisher’s exact test (one-tailed if not stated otherwise). Experimental data were averaged from at least three biological replicates and shown as box plots, with box bounds representing the first and the third quartiles and whiskers extending from the first and the third quartile to the lowest and highest data points or, if there are outliers, 1.5x of the interquartile range. Luciferase assays activity values were compared using the two-tailed Student’s t-test assuming unequal variances. Where necessary, p-values were adjusted for multiple testing using Benjamini–Hochberg correction. Specific tests used are indicated in the figures and/or figure legends.

## Data Availability

Research on the de-identified patient data used in this publication can be carried out in the Genomics England Research Environment subject to a collaborative agreement that adheres to patient led governance. All interested readers will be able to access the data in the same manner that the authors accessed the data. For more information about accessing the data, interested readers may contact or access the relevant information on the Genomics England website: https://www.genomicsengland.co.uk/research.

https://www.genomicsengland.co.uk/research

## Ethics statement

This work used data from the 100 000 Genomes Project (IRAS ID 166046; East of England - Cambridge South Research Ethics Committee reference 14/EE/1112), which obtained written informed consent from all participants (or from their parent/legal guardian). All investigations were conducted in accordance with the tenets of the Declaration of Helsinki.

This work was conducted under the registered and approved Genomics England Clinical Research project ID RR1011.

## Acknowledgements

This study has been possible thanks to a grant from Guy’s & St Thomas’ Charity grant STR111001.

This research was made possible through access to data in the National Genomic Research Library, which is managed by Genomics England Limited (a wholly owned company of the Department of Health and Social Care). The National Genomic Research Library holds data provided by patients and collected by the NHS as part of their care and data collected as part of their participation in research. The National Genomic Research Library is funded by the National Institute for Health Research and NHS England. The Wellcome Trust, Cancer Research UK and the Medical Research Council have also funded research infrastructure.

We thank Michael Simpson, Rebecca Oakey, Michael Antoniou, Anna Need, Suzi Walker, Eugene Makeyev, Vladimir Seplyarskiy, Patrick Campbell, Andriana Gialeli and Anna Zhuravskaya for insightful discussions, providing reagents and support.

## Data and code availability

Analysis of the 100,000 genomes project and NHS GMS data was performed inside the Genomics England Research Environment. We are happy to share the location of all code to registered users. Code used for analyses of external data outside Genomics England is available at GitHub: https://github.com/ykainov/PanelApp_genes.

---

**Supplementary Figures S1.**
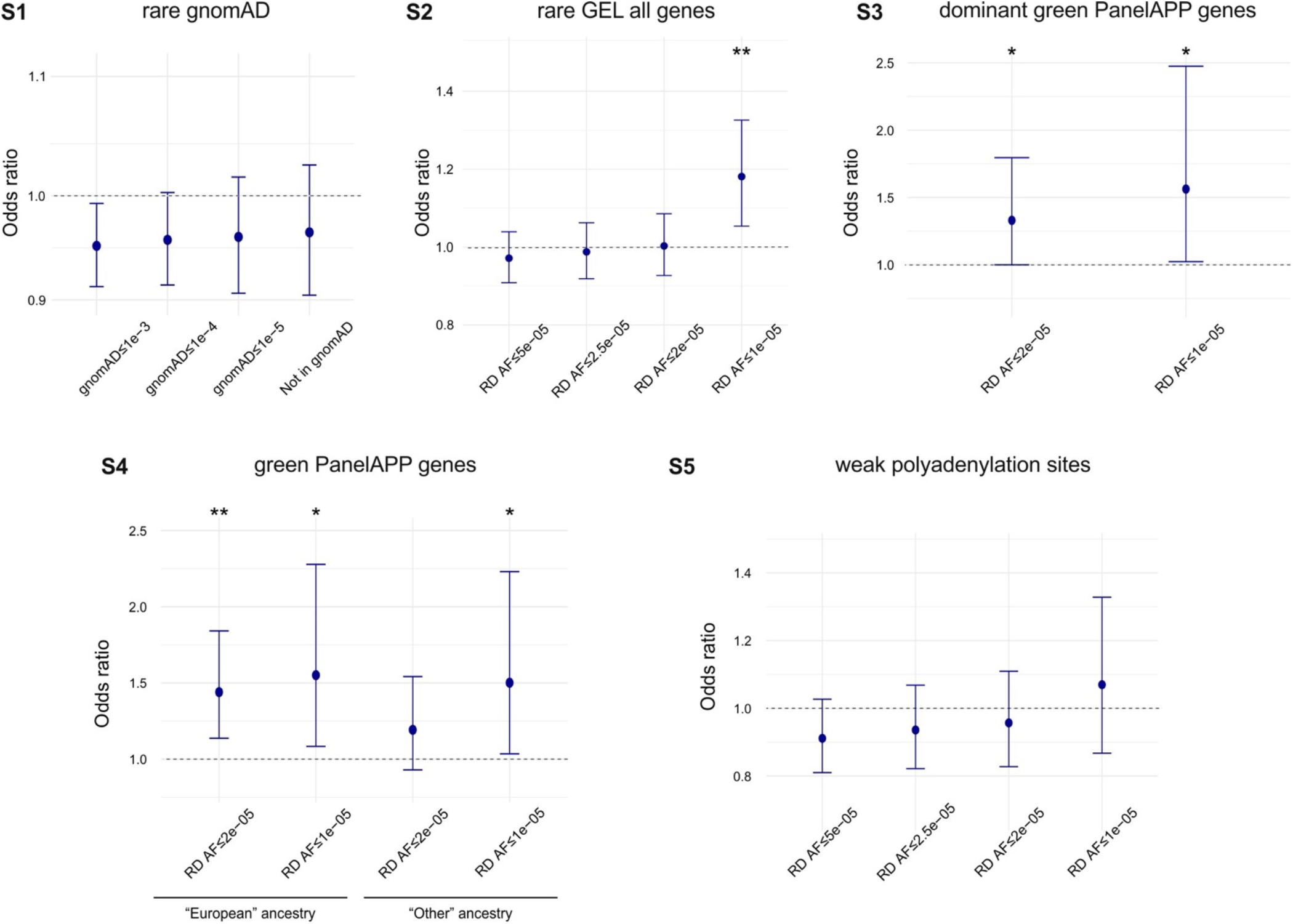
Odds ratios for an enrichment of rare gnomAD DOWNvars (gnomAD AF≤0.00005, GEL AF≤0.00005) in undiagnosed probands in comparison with diagnosed probands in all genes stratified by gnomAD AF. **S2 -** Odds ratios for an enrichment of rare DOWNvars (gnomAD AF=0, GEL AF≤0.00005) in undiagnosed probands in comparison with diagnosed probands in all genes stratified by GEL rare diseases AF. **S3 -** Odds ratios for the enrichment of rare DOWNvars (gnomAD AF=0, GEL AF≤0.00002) in undiagnosed probands in comparison with diagnosed probands in dominant green PanelApp genes. **S4 -** Odds ratios for the enrichment of rare DOWNvars (gnomAD AF=0, GEL AF≤0.00002) in undiagnosed probands of European inferred genetic ancestry (left) or any other inferred in comparison with corresponding diagnosed probands in green PanelApp genes. **S4 -** Odds ratios for the enrichment of rare DOWNvars (gnomAD AF=0, GEL AF≤0.00005) in weak polyadenylation sites in undiagnosed probands in comparison with diagnosed probands in green PanelApp genes stratified by GEL rare diseases AF. For all panels whiskers represent 95% confidence intervals, ** - BH-adjusted one-tailed Fisher’s exact test p-value < 0.01, * - BH-adjusted one-tailed Fisher’s exact test p-value < 0.05

**Table S1.**
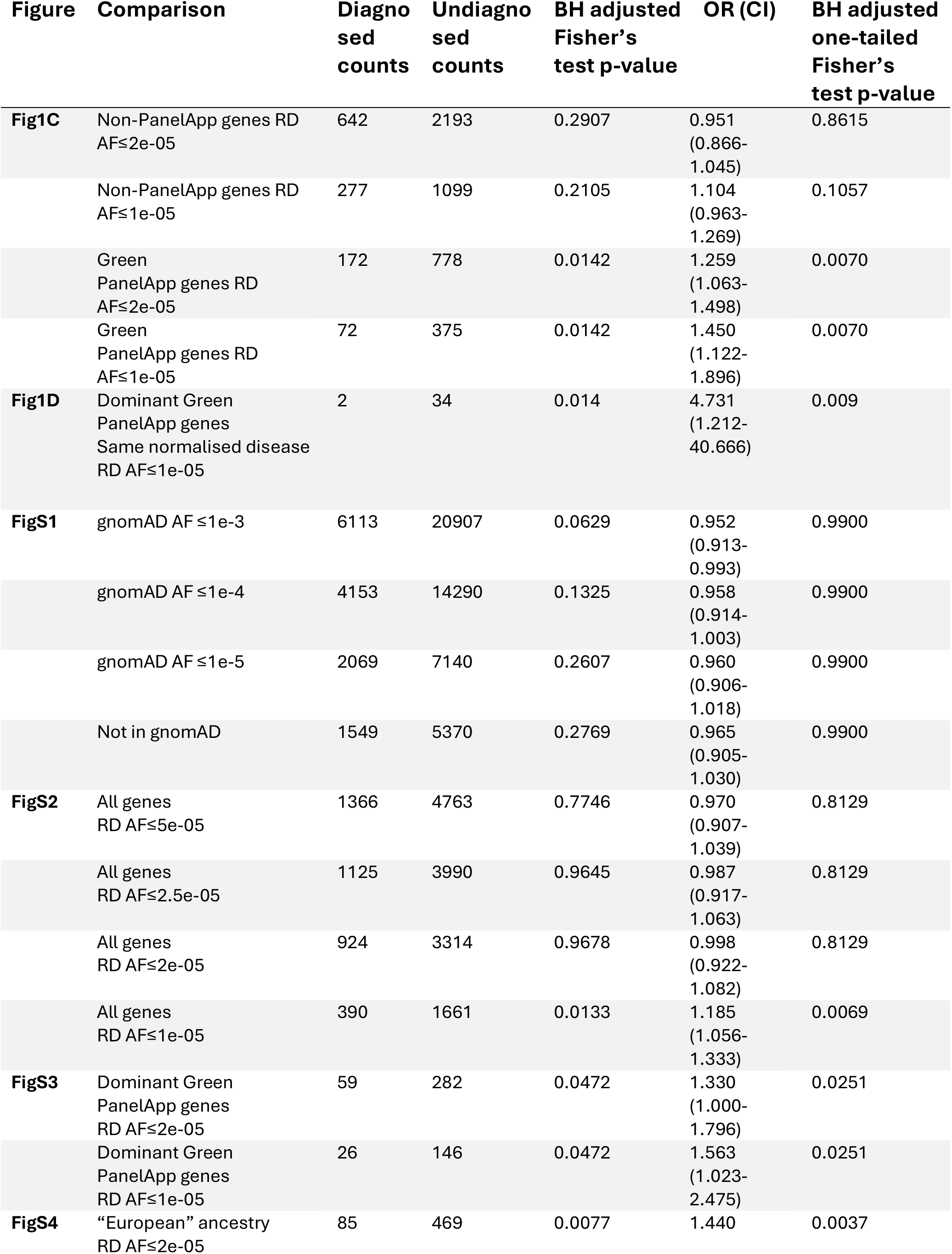
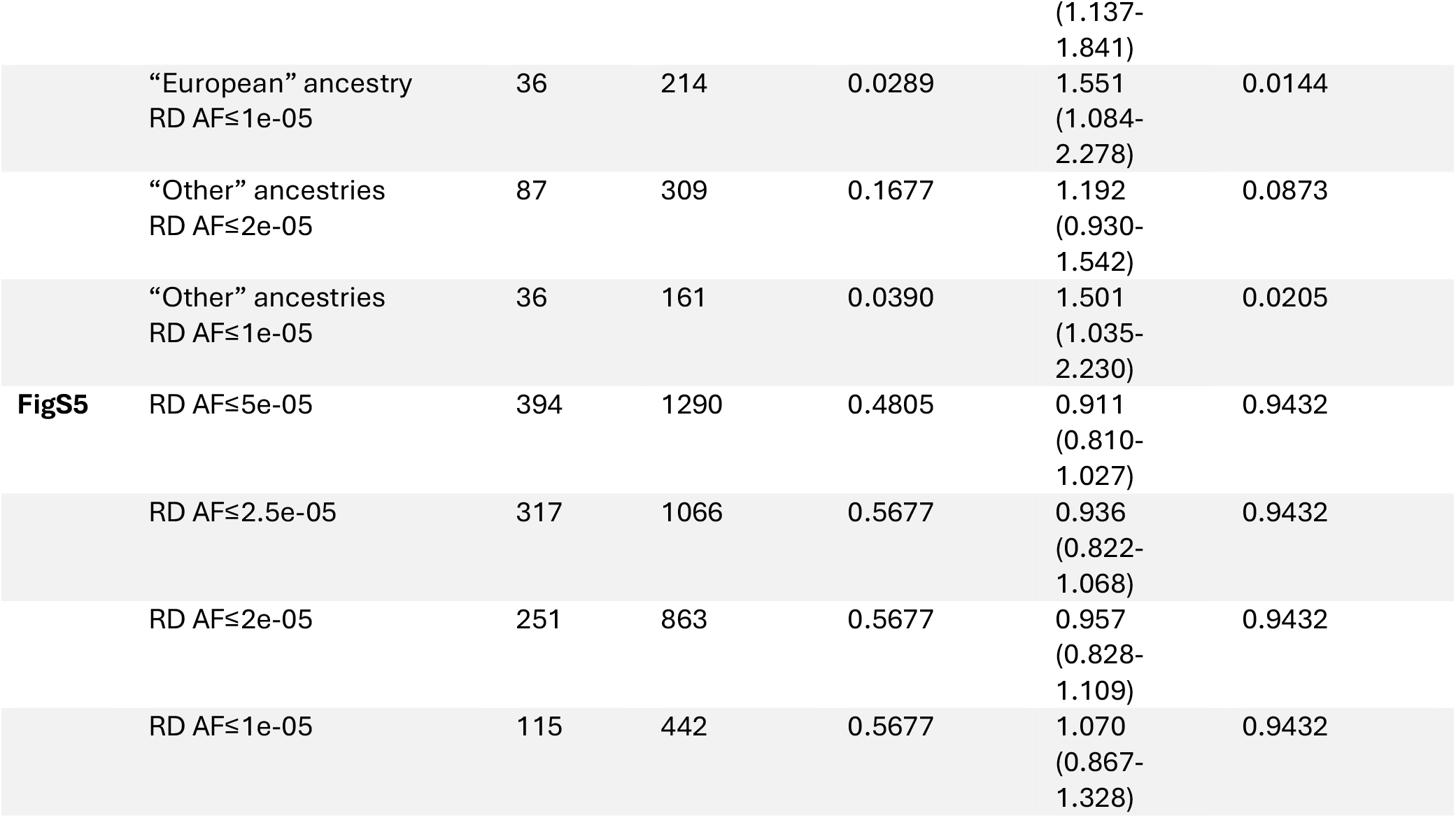
Burden testing statistics.

